# Adversity and brainAGE associations with age of first cannabis use in the Adolescent Brain Cognitive Development (ABCD) Study

**DOI:** 10.64898/2026.08.18.26360711

**Authors:** Karina Aika Thiessen, Yuetong Yu, Laura Schmid, Alexis Brieant, Sophia Frangou, Christian Georg Schütz

**Author notes:** **Corresponding author:** Karina Aika Thiessen, 430 - 5950 University Blvd, Vancouver, BC Canada V6T 1Z3.

## Abstract

**Importance:** Adolescent cannabis use is a growing concern due to its associations with long-term adverse mental health outcomes. However, the distinct, temporal associations of neurodevelopmental factors and childhood adverse life experiences (ALEs) with adolescent substance use in have not yet been fully elucidated. The Adolescent Brain Cognitive Development (ABCD) Study offers an unprecedented opportunity to prospectively examine neurobiological and socioenvironmental predictors of cannabis onset.

**Objective:** To investigate magnetic resonance imaging-derived neurodevelopmental cortical brain Age Gap Estimate (brainAGE) and adverse life events as risk factors of early cannabis initiation.

**Design, Setting, and Participants:** The ABCD Study is a longitudinal study across 22 sites in the United States. Data are collected starting at approximately 10 years old (currently at year-7 follow-up). Our analyses comprised 6688 (48% female) youth after exclusions.

**Main Outcomes and Measures:** Cox proportional hazard models were computed to investigate brainAGE-sex interactions and 10 adversity dimensions at baseline as predictors of time to cannabis initiation up to age 18.

**Results:** Mean age of initiation was 14.8 years (SD=1.43). Global brainAGE was modestly associated with cannabis initiation in females only (Hazard Ratio [HR]=1.05; 95% Confidence Interval [CI]=1.00-1.10, *p* = .049). Low socioeconomic status, caregiver substance use, family anger and arguments, and caregiver lack of supervision were associated with initiation (HRs = 1.11, 1.56, 1.09, 0.85, respectively; CIs = 1.04-1.19, 1.44-1.69, 1.09-1.19, 0.76-0.91, respectively; *p*’s < .05). Other ALE dimensions and network-specific brainAGEs were not significantly associated with initiation.

**Conclusions and Relevance:** Findings suggest that while cortical brainAGE may be somewhat increase vulnerability to adolescent cannabis use in females, its contributions are modest at best. In contrast, early childhood adversities such as socioeconomic factors and familial characteristics may present more substantive targets for prevention and intervention.

**KEY POINTS:** *Question:* What is the relationship between neurobiological brainAGE and early life adversity in childhood to subsequent adolescent cannabis use?

*Findings:* BrainAGE was prospectively associated with cannabis onset in females, but effect sizes were limited. Specific early life adversities were the strongest predictors of cannabis initiation.

*Meaning:* Early substance use preventive strategies may particularly benefit from targeting socioenvironmental and familial factors.

## 1. Introduction

Cannabis is among the most widely used substances globally^1^. In the United States, cannabis use disorder is one of the most common substance use disorders^2^. Adolescent cannabis use may be an important modifiable risk factor for prevention of substance use disorders and other adverse outcomes^3^. For example, a twin study found that those who used cannabis by age 17 were up to five times more likely to use other substances or have a substance use disorder^4^. More broadly, adolescent cannabis use and younger age of first use have been found to predict more substance use disorder symptoms in adulthood^5^, earlier onset of psychosis^6,7^, decreases in neurocognitive functioning^8^, and reduced academic achievement^8^. Adolescent substance use is also associated with increased risk for other psychiatric disorders and relational, academic, and employment difficulties^9^.

Exposure to adverse life experiences (ALEs) during childhood and adolescence has been widely linked to substance-related problems^10–12^, suggesting potential preventive targets. However, ALEs encompass a wide range of experience across domains, including physical, emotional, and sexual abuse; physical and emotional neglect; caregiver mental health and substance use; and socioeconomic status. Using factor analysis, Brieant et al.^13^ previously identified 10 dimensions of ALEs across over 11,000 youth in the Adolescent Brain Cognitive Development (ABCD) Study. This data-reduction approach allows for the examination of the unique contribution of different dimensions of ALEs in terms of risk for early adolescent substance use.

Further, early substance initiation and related outcomes may be influenced by neurobiological characteristics^14–16^. Recent literature has identified differences in gray matter volume, particularly thinner fronto-cortical regions, at age 9-10 years old among those that used substances by age 15 in the ABCD Study^15^. These differences in gray matter volume may reflect differences in neurodevelopmental trajectories, with increased neuroplasticity, cortical synaptic pruning, decreases in gray matter, and increases in white matter occurring during adolescence^16–20^. In particular, brain networks involved in cognitive control and emotion regulation – both processes implicated in higher-risk substance use – undergo substantive changes during adolescence^14,21^. Corresponding increases in risk-taking behavior and motivational salience occur throughout adolescence, relative to both childhood and adulthood^14,18,21^. Neurobiological factors associated with substance use may also differ by sex^22^.

To characterize potential neurodevelopmental trajectories, machine learning can be applied to estimate biological brain age from structural magnetic resonance imaging (MRI), with the difference between estimated age and chronological age termed the Brain Age Gap Estimate (brainAGE)^23,24^. In youth, higher brainAGE has been linked to stronger cognitive performance, though associations with health-risk behaviors, such as substance use, are less consistent^25,26^. BrainAGE has not yet been examined in relation to adolescent substance use but demonstrates potential as a quantitative marker of neurodevelopmental deviation.

Given the extensive neurodevelopmental changes in adolescence, the potential role of neurodevelopmental trajectories and environmental factors on cannabis use onset must be prospectively examined to identify targets for prevention and early intervention^27,28^. The ongoing ABCD Study follows youth from 9-10 years old for 10 years, collecting data on mental health, substance use, caregiver characteristics, socio-environmental factors, genetics, and neuroimaging-based measures. With its large, diverse cohort and longitudinal design, the ABCD Study offers an unparalleled opportunity to investigate the role of environmental and neurobiological factors over time with exceptional scientific rigor.

In this paper, we investigated the associations of ALEs and global brainAGE with time to first cannabis use in the ABCD Study. We hypothesized that ALEs and brainAGE would be uniquely associated with age of first cannabis use. Given prior literature identifying sex differences in neurobiological risk for adolescent substance use^22^, we also considered sex-brainAGE interactions. Beyond the established global measure of brainAGE, we introduce network-level brainAGE^29^, a novel approach that incorporates spatial variation in age-related changes across brain networks and may reveal more fine-grained networks relevant to adolescent substance use. We conducted exploratory analyses for the control, salience, and limbic networks^30^.

## 2. Methods

This analysis utilizes data from the Adolescent Brain Cognitive Development (ABCD) Study, a 10-year longitudinal cohort study following youth across 22 sites in the United States. Baseline assessments were conducted at approximately 9-10 years old and cannabis use data was collected biannually. As the ABCD Study is ongoing, we utilize the most recent cannabis use data up to age 18. In the cases of siblings, one member of each family was randomly selected and included in our analyses to maintain independence of observations. Participants that had missing data or whose T1-weighted (T1W) structural MRI scans did not meet quality control standards from ABCD Study recommendations^31^ or during image processing. Participants that reported prior cannabis initiation at baseline were also excluded from all analyses. See supplementary materials (S1 eFigure 1) for the participant inclusion/exclusion flow diagram.

### 2.1. Adverse Life Experience (ALE) Scores

Given the range of potential ALEs to consider, we used previously-determined baseline ALE factor scores^13^. Factors were derived from 139 potential adversity variables that were then aggregated into 60 variables based on intercollinearity, resulting in 10 emergent factors based on caregiver- and youth-reports (CGR and YR, respectively). The 10 factors were: (1) CGR caregiver psychopathology, (2) CGR socioeconomic disadvantage and lack of neighborhood safety, (3) YR primary caregiver lack of support, (4) YR secondary caregiver lack of support, (5) YR family conflict, (6) CGR caregiver substance use/separation from biological parent, (7) CGR family anger and arguments, (8) CGR familial physical/emotional aggression, (9) CGR physical trauma exposure, and (10) YR caregiver lack of supervision. Each participant received a standardized score for each of the 10 dimensions, with higher scores indicating more adversity. See Brieant et al. for additional factor analysis methods^13^.

### 2.2. Computation of BrainAGE Measures

Baseline T1-weighted MRI data were obtained from the ABCD Study repository held in the NIMH Data Archive. ABCD structural MRI data were acquired using a harmonized multi-site protocol implemented on Siemens, GE, and Philips scanners, including high-resolution 3D T1-weighted magnetization-prepared rapid acquisition gradient echo images for cortical and subcortical segmentation, as described previously^32,33^. Images from ABCD data release 5.1 were processed locally at the University of British Columbia using FreeSurfer Version 7.1 using standard pipelines and cortical thickness and surface area measures were extracted for estimating sex-specific brainAGE measures. Global brainAGE was estimated using a previously published developmental brain-age model while network brainAGE was estimated using a separate model, both available through the CentileBrain platform (https://centilebrain.org/)^24,29^. Additional details on brainAGE calculations, model development, and validation can be found in supplementary materials (S2).

For all brainAGE measures, the gap estimate was calculated as the neuroimaging predicted brain age minus chronological age, with positive values indicating higher-than-expected brain age and negative values indicating lower-than-expected brain age.

### 2.3. Cannabis Use

Cannabis use data were collected biannually via youth self-report^34^. As over 50% of participants had not yet completed year 7.5 substance use phone interviews at the time of analysis, only cannabis use data up to year 7.0 follow-up were utilized. Time to first cannabis use was extracted from summary substance use data provided within the ABCD study data release^31^. Whether or not a participant engaged in any cannabis use was coded as binary variables (yes/no) based on tabulated summary data.

### 2.4. Analysis: Cox Proportional Hazard Models

Mixed effects Cox proportional hazard models were computed to evaluate the relationships between ALEs, brainAGE, and cannabis initiation. Continuous predictors were 99% winsorized. To account for left truncation with baseline occurring at approximately age 10 years old, age at study entry was input into the models as participant start time. Stop time was age of first cannabis use, or age at most recent follow-up in the case of censored outcomes (i.e., no reported cannabis use). Cannabis initiation was coded as a binary outcome to indicate event occurrence. Our primary multivariate model included sex, the 10 ALEs, and global brainAGE. Main effects and interactions between sex assigned at birth and brainAGE were examined. We also conducted exploratory network-specific brainAGE analyses. Scanner site (random effect) and age at baseline as were included as covariates in all models. Race and ethnicity were not included in our models according to prior recommendations^35^ and ABCD responsible use of data guidelines^31^ and pubertal stage was excluded to prevent potential confounding with brainAGE^36^.

Assumptions of linearity of log-hazards and no influential observations were tested via visual inspection and computation of difference of betas, respectively. Assumptions of proportional hazards for ALEs except secondary and primary caregiver lack of support and familial anger and arguments were violated when proportional hazards tests on Schoenfeld residuals were conducted (*p*’s < .05). However, this violation was deemed minor upon visual inspection, and prior literature has found that effects of such violations are minimal^37^. Given the large sample size, effect sizes and confidence intervals are prioritized in interpretation of findings, beyond nominally reported *p*-values.

## 3. Results

### 3.1 Demographics and Descriptives

Our analyses comprised a sample of *n* = 6688 (52% male) out of the original ABCD Study sample of *N =* 11,875. Of the included sample, 25% reported cannabis initiation.

Mean self-reported age of initiation was 14.8 (SD = 1.43) years old. Additional descriptive statistics are detailed in Table 1. There were no significant differences in cannabis initiation between participants included in our analyses and participants excluded due to missing data (Hazard Ratio [HR] = 1.01, 95% Confidence Interval [CI] = 0.92-1.10, *p* = 0.822).

**Table 1.** Descriptive statistics by cannabis use status.

|  | No initiation<br>N (%) or Mean (SD) | Initiation<br>N (%) or Mean (SD) | Total<br>N (%) or Mean (SD) |
| --- | --- | --- | --- |
| *n* | 5048 (75) | 1640 (25) | 6688 (100) |
| Age at baseline (years) | 9.91 (0.62) | 10.08 (0.60) | 9.95 (0.62) |
| Sex assigned at birth |  |  |  |
| Female | 2384 (47) | 812 (50) | 3,196 (48) |
| Male | 2664 (53) | 828 (50) | 3,492 (52) |
| Age of cannabis initiation (years) | N/A | 14.8 (1.43) | N/A |
| Caregiver psychopathology (CGR) | -0.03 (0.90) | 0.11 (0.99) | 0.00 (0.93) |
| Socioeconomic disadvantage and lack of neighborhood safety (CGR) | -0.03 (0.88) | 0.12 (0.93) | 0.01 (0.89) |
| Secondary caregiver lack of support (YR) | 0.07 (0.77) | 0.08 (0.76) | 0.07 (0.77) |
| Primary caregiver lack of support (YR) | 0.07 (0.77) | 0.07 (0.76) | 0.07 (0.77) |
| Family conflict (YR) | 0.06 (0.76) | 0.06 (0.76) | 0.06 (0.76) |
| Caregiver substance use/separation from biological parent (CGR) | 0.04 (0.62) | 0.29 (0.73) | 0.10 (0.66) |
| Family anger/arguments (CGR) | 0.00 (0.73) | 0.11 (0.75) | 0.02 (0.73) |
| Family physical/emotional aggression (CGR) | 0.06 (0.69) | 0.15 (0.73) | 0.08 (0.70) |
| Physical trauma exposure (CGR) | 0.04 (0.58) | 0.16 (0.63) | 0.07 (0.59) |
| Caregiver lack of supervision (YR) | 0.02 (0.74) | -0.09 (0.67) | -0.01 (0.73) |
| Global brainAGE | -0.78 (1.60) | -0.75 (1.71) | -0.78 (1.63) |
| Limbic network brainAGE | -0.86 (1.80) | -0.83 (1.91) | -0.85 (1.83) |
| Control network brainAGE | -1.00 (1.69) | -0.97 (1.84) | -0.99 (1.73) |
| Salience/ventral attention network brainAGE | -0.92 (1.77) | -0.87 (1.88) | -0.91 (1.80) |

Note: brainAGE = brain Age Gap Estimate. CG = caregiver reported. YR = youth reported. Higher adverse life event factor scores indicate more adversity.

### 3.2. Primary Analysis: Global brainAGE and ALEs

Results of the primary analysis with global brainAGE, including hazard ratios and *p*-values are shown in Figure 1 respectively. The overall regression fits were significant (log likelihood [LR] *χ*^2^ = 355.0 on 15 degrees of freedom [*df*], *p* < .001).

**Figure 1.**
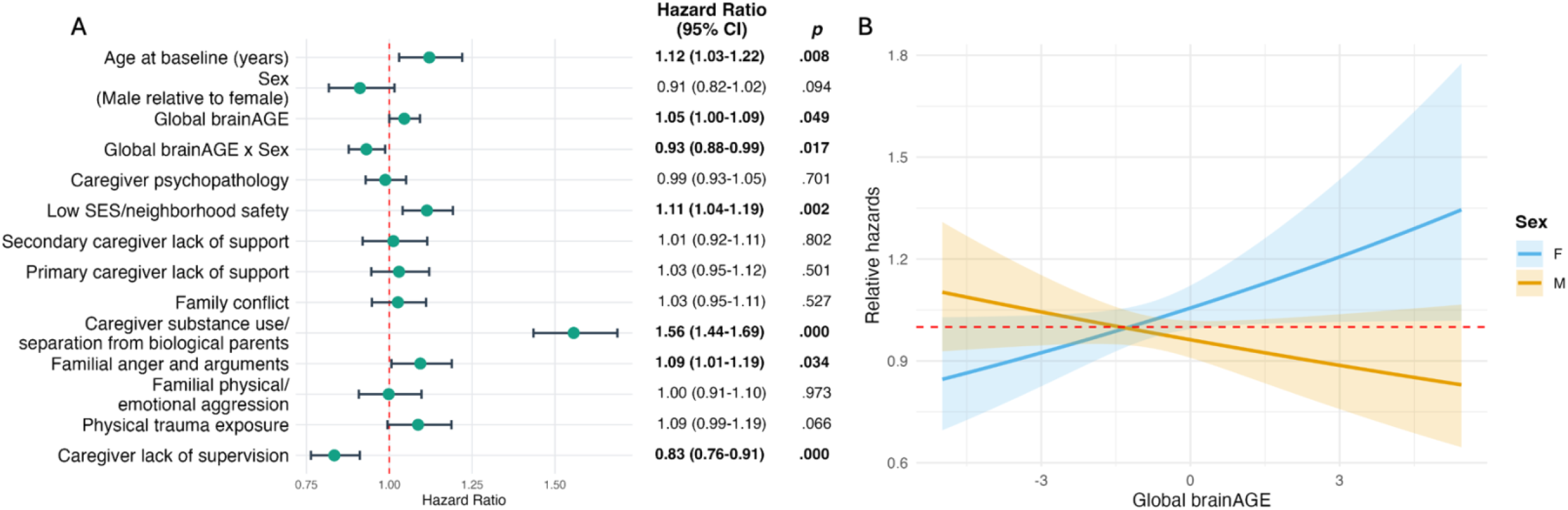
Results of global brainAGE primary analysis and post-hoc interaction test. Some statistically significant findings appear to include lower limits of confidence intervals at 1.00 due to rounding. **(A)** Hazard ratios of cannabis initiation by sex, ALEs, and global brain Age Gap Estimate (brainAGE). Error bars indicate 95% CI. Log likelihood *χ*^2^ = 355.0 on 15 degrees of freedom, *p* < .001. Statistically significant findings are indicated in bold. **(B)** Relative hazards of brain Age Gap Estimate (brainAGE) by sex interactions. Shading indicates 95% confidence intervals (CI). F = female. M = Male. HR_F_ = 1.045 (1.00-1.10, *p* = .049), HR_M_ = 0.973 (0.94-1.01, *p* = .176), interaction *p* < .017.

After controlling for scanner and age at baseline, we identified a significant interaction between sex and global brainAGE (Figure 2; *p* = .017). Post-hoc tests identified a small but significant association between global brainAGE and cannabis initiation in females but not males (females: HR = 1.045, 95% CI = 1.00-1.10, *p* = .049; males: HR = 0.973, 95% CI = 0.94-1.01, *p* = .176). Low SES/neighborhood safety, caregiver substance use/separation from biological parents, and family anger and arguments, had higher hazards of initiation (HR = 1.11, 1.56, 1.09, respectively; CI = 1.04-1.19, 1.44-1.69, 1.09-1.19, respectively; *p*’s < .05). In contrast, caregiver lack of supervision was associated with lower hazards of initiation (HR = 0.85, CI = 0.76-0.91, *p <* .001). Other ALEs were not significantly associated with cannabis EXP. Findings were largely the same for initiation.

ALE findings were overall stable across global, limbic, control, and salience network brainAGE models. There were no other significant brainAGE or sex-specific findings in the network-specific models. Exploratory network-specific brainAGE model results are reported in supplementary materials (S3 eFigures 2-4).

## 4. Discussion

By leveraging a large, longitudinal dataset, we were able to prospectively examine cannabis onset across multiple dimensions of ALEs and developmental brainAGE in a diverse sample. We examined relationships between age of cannabis initiation up to age 17-18 years old, sex-brainAGE interactions, and specific early childhood ALEs. Cannabis initiation was relatively common among participants.

The relationship between the cumulative childhood ALEs and substance use is well-characterized^11^, but it is less understood how individual adversities may uniquely contribute to risk. By utilizing data-driven ALE factor scores, we were able to empirically differentiate between ALE types that may be prioritized as targets for intervention. The relationship between SES/neighborhood characteristics and cannabis use has been established in prior literature and was replicated in our analysis^38–41^.

Caregiver substance use and separation from biological parents factor scores were associated with increased likelihood of cannabis initiation, as is consistent with prior substance use literature^12,42,43^, and had the largest effect size out of the variables of interest. While further research will be needed to elucidate mediating factors, we propose that this relationship may be due to several reasons, such as increased access to substances, social learning from parents^44^, or heritability of substance use behaviors^45^.

Previous literature has also identified relationships between non-health-related parent-child separation and child risk for future SUDs^46^. Separation from biological parents may disrupt parent-child relationships, which may be related to maladaptive coping strategies. Thus, children with parents with substance-use related problems may be at higher risk for early cannabis use, and both parents and children may also benefit from targeted early prevention strategies.

Notably, there was variability in the associations between familial interpersonal dynamics and cannabis use. Findings differed by nature of the familial challenges: caregiver-reported family anger/arguments dimension was associated with cannabis initiation while caregiver reported physical/emotional aggression was not. Further, the youth-reported family conflict dimension was not significantly associated with cannabis initiation. Perception of ALEs may vary by reporting source or there may be differences in reliability of reporting between caregivers and children. Prior studies have found limited agreement between child and caregiver reports of maltreatment, with evidence suggesting caregiver underreporting but variation in concordance by child age at time of reporting and form of maltreatment^47,48^.

Physical trauma exposure – including physical and sexual assault – was unassociated with cannabis outcomes in our analyses. This may similarly be due to underreporting of childhood maltreatment. Alternatively, a review found sex-specific relationships between physical abuse and substance use problems, with physical trauma generally associated with substance use problems in females but not males^49^; further investigation of sex differences may be warranted.

Youth-reported caregiver lack of supervision *decreased* hazards of cannabis initiation. Prior literature including a meta-analysis and a study amongst a subsample of ABCD study participants has found higher parental monitoring to be protective^50,51^. It is unclear why our findings using factor scores may be different but may reflect differences in reporting and scoring methodology.

Sex and global brainAGE interactions suggest that neurodevelopmental trajectories may have small but distinct associations with cannabis use phenotypes in females relative to males. Specifically, higher global brainAGE slightly increased hazards of cannabis EXP and initiation in female participants but was not significantly associated with cannabis outcomes in males. BrainAGE estimates were computed separately for males and females (S1), so this relationship was not due to sex-based biases in computed brainAGE. A prior review suggested that sex differences in adolescent vulnerability for substance use may stem from differences in motivational pathways^22^. While our data supports the possibility of brainAGE differences to some extent, findings should be interpreted with caution due to small effect sizes in females for global brainAGE analyses. While we did not find network-specific main effects or interactions, we looked at lower-threshold cannabis use patterns, and it is possible that network-specific findings may emerge with higher-threshold or escalating cannabis use.

Taken together, our findings suggest that the potential neurodevelopmental factors in cannabis risk may vary by sex, but potential associations are modest. Further, some ALEs may be more relevant than others in risk of early adolescent cannabis use and should be prioritized as targets for intervention. Reports of ALEs may also vary by reporting source (i.e., child/youth vs caregiver). Our findings further suggest that ALEs occurring in childhood may have longer-term impacts on adolescent behavior, and early initiation of preventive interventions focused on socioeconomic factors and familial dynamics may have benefits. SES- and family-based interventions that focus on caregiver behaviors and family dynamics may be particularly important.

This study has some limitations. First, while we examined cannabis use prospectively, etiology cannot be determined due to the observational nature of the study. Interpretation of findings should be taken with caution, and causal pathways should not be assumed. Second, the organization of Yeo’s networks and their function have been debated^52,53^ and there is heterogeneity across individuals^54^. Nonetheless, the relationship between substance use and the networks we examined are well-characterized. Third, there may be cohort effects (e.g., due to the COVID-19 pandemic) and sampling effects (e.g., study attrition) that limit generalizability of findings^55,56^. However, we found no significant differences in cannabis initiation when comparing participants included in our analyses versus those excluded due to missingness. Lastly, we examined relatively low-threshold cannabis use. Due to the young age of our sample and the established relationship between early adolescent cannabis use and later neurocognitive, psychiatric, and social challenges, this metric remains meaningful. However, we anticipate that cannabis use will increase with age in the study cohort and further investigations of biopsychosocial factors in relation to escalating cannabis use will be warranted.

To our knowledge, this is the first study examining brainAGE and ALE dimensions as risk factors for early adolescent cannabis use in the ABCD cohort. We identified distinct associations between cannabis use outcomes and specific forms of ALEs, as well as sex-brainAGE interactions. By utilizing brainAGE, we were able to explore potential relationships between individual differences in neurodevelopmental trajectories and adolescent cannabis use, compared to conventional morphological studies. Future studies may examine risk factors in consideration of higher threshold cannabis use later in adolescence or early adulthood.

## Data Availability

All data can be requested from the ABCD Study NBCD data archive (https://www.nbdc-datahub.org)

https://abcdstudy.org

https://www.nbdc-datahub.org

https://centilebrain.org/#/

## Data Source and Access

Data used in the preparation of this article were obtained from the Adolescent Brain Cognitive Development™ (ABCD) Study, held in the NIH Brain Development Cohorts Data Sharing Platform. This is a multisite, longitudinal study designed to recruit more than 10,000 children aged 9–10 and follow them over 10 years into early adulthood.

The ABCD Study® is supported by the **National Institutes of Health** and additional federal partners under award numbers:

U01DA041048, U01DA050989, U01DA051016, U01DA041022, U01DA051018, U01DA051037, U01DA050987, U01DA041174, U01DA041106, U01DA041117, U01DA041028, U01DA041134, U01DA050988, U01DA051039, U01DA041156, U01DA041025, U01DA041120, U01DA051038, U01DA041148, U01DA041093, U01DA041089, U24DA041123, U24DA04114 7. A full list of supporters is available at [Federal Partners – ABCD Study](https://abcdstudy.org/federal-partners.html).

ABCD Consortium investigators designed and implemented the study and/or provided data but did not necessarily participate in the analysis or writing of this report. This manuscript reflects the views of the authors and may not reflect the opinions or views of the NIH or ABCD Consortium investigators. The ABCD data repository grows and changes over time. Adversity data came from doi: 10.15154/cqdy-5453. ABCD neuroimaging data came from 10.15154/z563-zd24. All other data used in this report came from doi: https://doi.org/10.82525/8f3w-5260.

Google Gemini Version 3 was used to edit code to create tables and figures and for reviewing text content.

## Author contributions

KAT: conceptualization, methodology, data curation, formal analysis, visualization, writing – original draft, writing – review and editing, project administration

YY: data curation, methodology, formal analysis, writing – review and editing LS: conceptualization, writing – review and editing

AB: data curation, methodology, formal analysis, writing – review and editing

SF: conceptualization, methodology, writing – review and editing, supervision

CS: conceptualization, methodology, writing – review and editing, supervision

