## Supplementary materials for "Adversity and brainAGE associations with age of first cannabis use in the Adolescent Brain Cognitive Development (ABCD) Study"

#### *S1. Inclusion/Exclusion Flow Diagram*

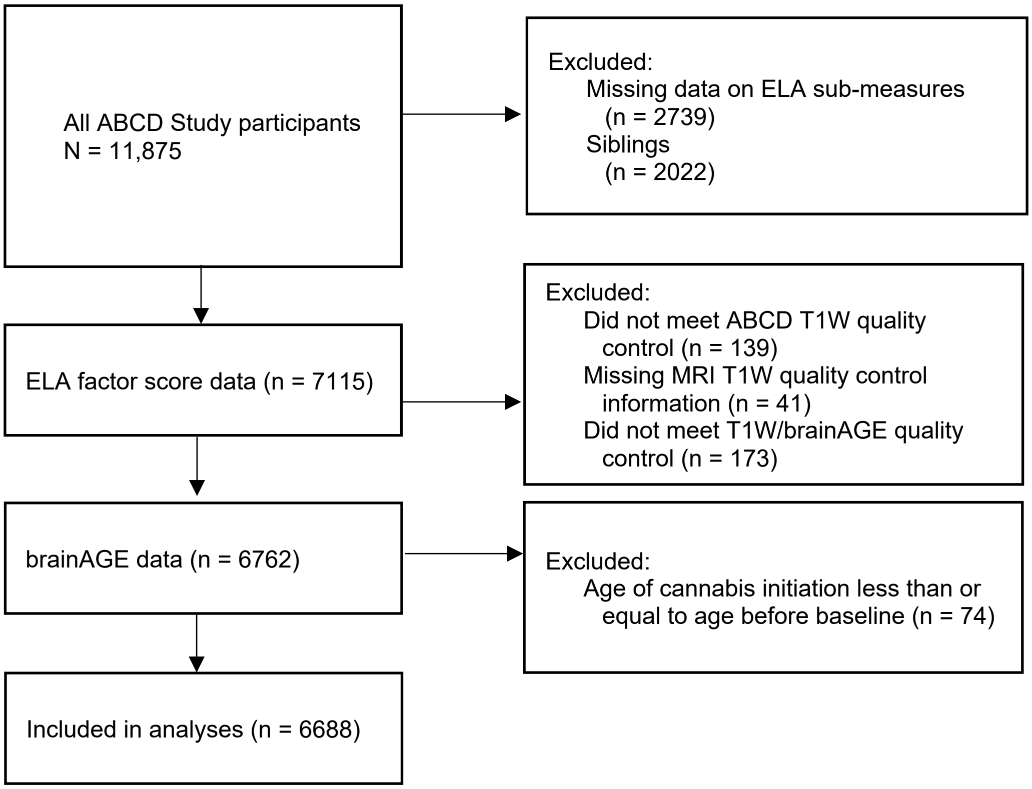


**eFigure 1. Inclusion/exclusion flow diagram**

#### *S2. BrainAGE Model Development and Validation*

The developmental global brainAGE model available through the CentileBrain platform (https://centilebrain.org/) was developed using structural MRI data from 2,105 typically developing individuals aged 5 to 22 years from five cohorts[24]. The global brainAGE model used sex-specific support vector regression with a radial basis function kernel and cortical thickness and surface area features from the Schaefer 400-parcel atlas. Pretrained model parameters were applied to the corresponding Schaefer 400-parcel cortical features extracted from the current ABCD sample.

The network brainAGE model, available through the CentileBrain platform (https://centilebrain.org/, was trained in 9,473 healthy individuals aged 3 to 92 years[29]. Cortical thickness and surface area features were extracted using the Schaefer 1,000-parcel atlas. Each parcel was assigned to one of the seven Yeo functional networks[30]— default mode, dorsal attention, ventral attention/salience, frontoparietal control, limbic, somatomotor, and visual — based on maximal spatial overlap, defined as the greatest number of shared vertices between each Schaefer parcel and the Yeo 7-network atlas. Sex-specific support vector regression models with radial basis function kernels were then applied to estimate network-specific brainAGE. For the current study, pretrained model parameters developed in individuals younger than 40 years were applied to the corresponding Schaefer 400-parcel cortical features extracted from the current ABCD sample.

### *S3. Network-specific brainAGE models*

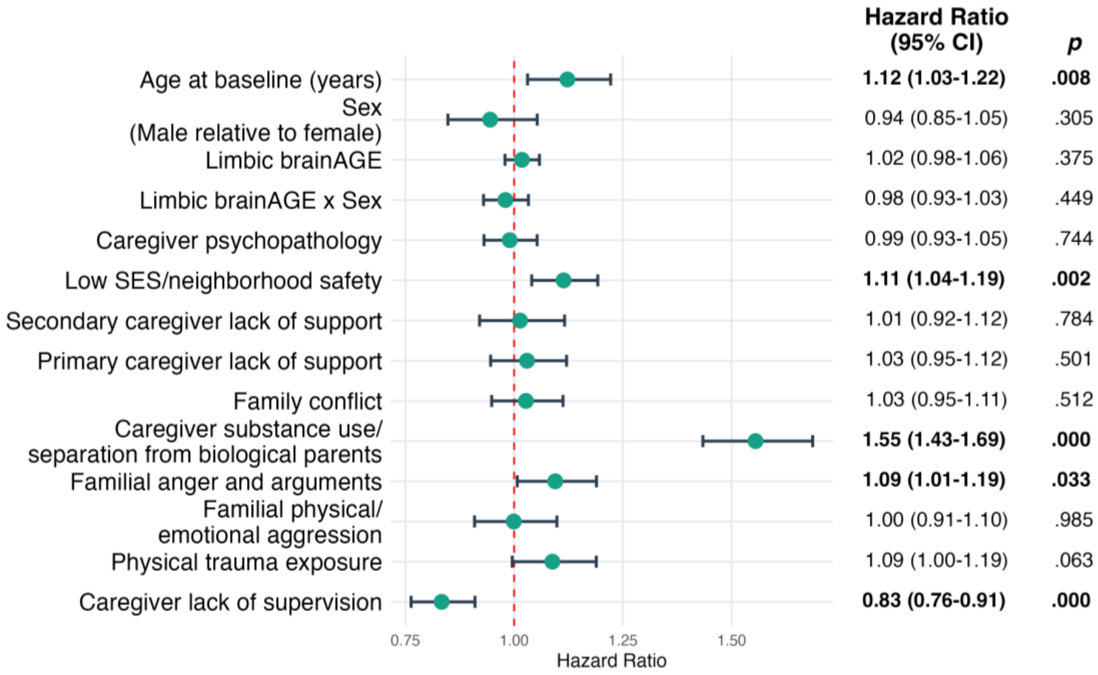


**eFigure 2. Cannabis initiation – limbic network brainAGE model.**

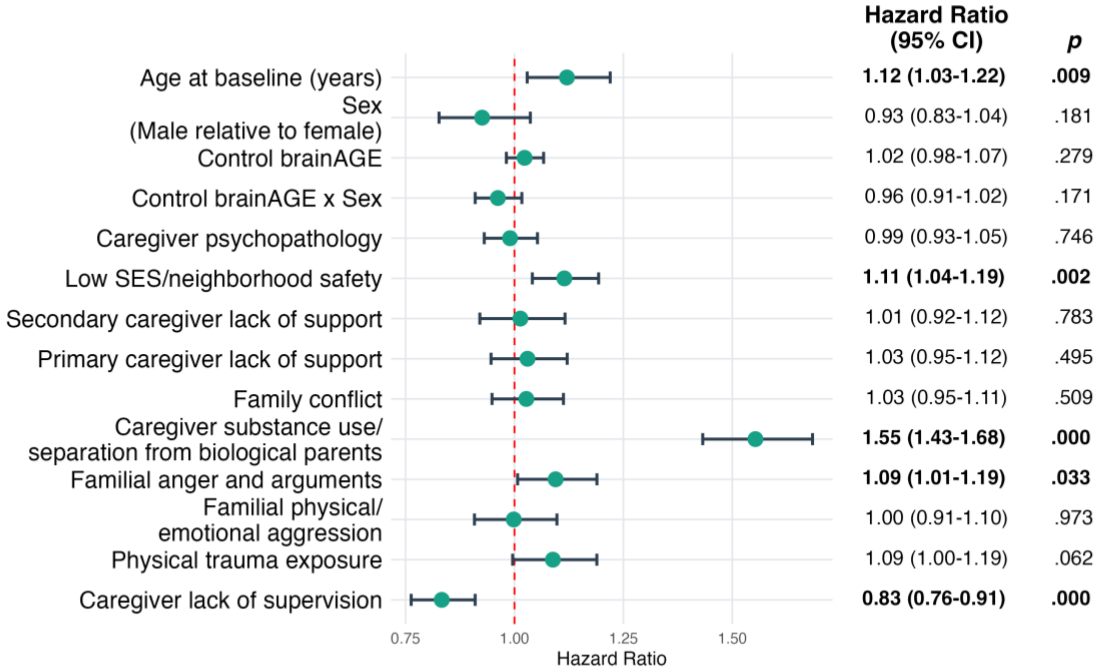


**eFigure 3. Cannabis initiation – control network brainAGE model.**

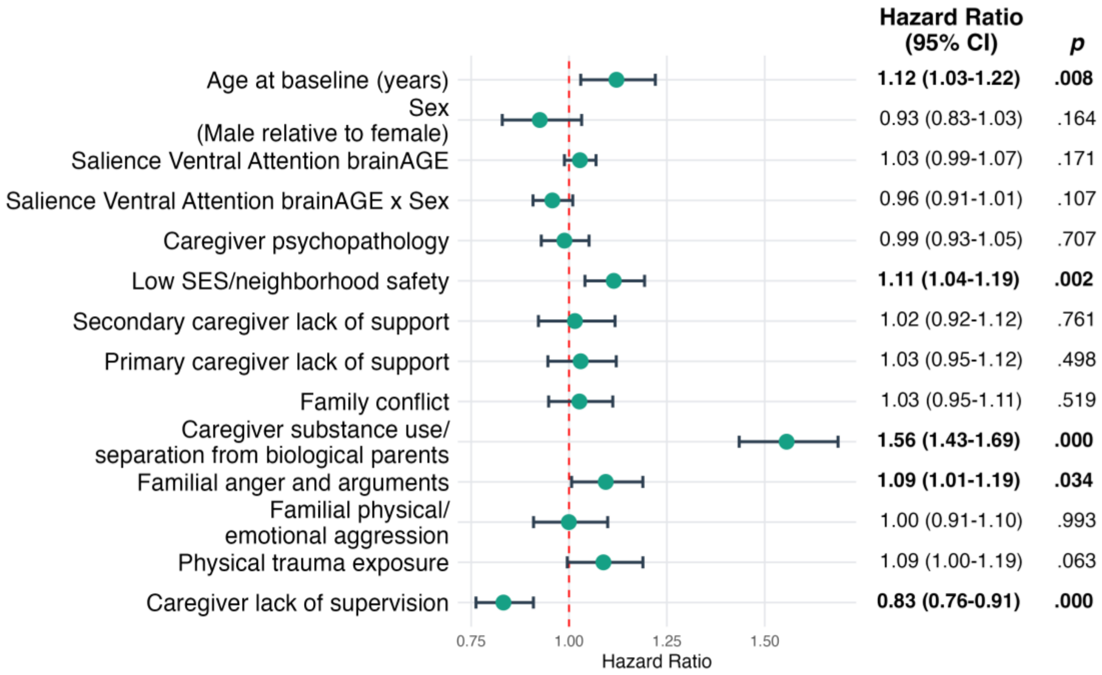


**eFigure 4. Cannabis initiation – salience/ventral attention network brainAGE model.**
